# Knowledge and perceptions on COVID-19 among Senior High School students in Ghana: a cross-sectional study

**DOI:** 10.1101/2020.11.18.20234088

**Authors:** Isaac Bador Kamal Lettor, Paschal Awingura Apanga, Maxwell Tii Kumbeni, Ramatu Akunvane, Robert Akparibo

## Abstract

**Background:** The COVID-19 pandemic is associated with high morbidity and mortality. In Ghana, policy interventions have been implemented by the Government to combat the pandemic. However, the knowledge and perceptions of Senior High School students are not investigated on the COVID-19 symptoms, transmission and the government policy measures.

**Objectives:** The present study surveyed senior high school students to assess their knowledge and perceptions of COVID-19 and the government policy measures to address the outbreak.

**Methods:** The study employed a descriptive cross-sectional study design to assess the knowledge and perceptions of senior high school students on the COVID-19 pandemic and the measures put in place to address it. 624 senior high school students aged 18 years old and above were surveyed. Descriptive analysis was performed to assess knowledge and perceptions of COVID-19 symptoms, mode of transmissions and prevention.

**Findings:** Most students were knowledgeable about COVID-19 symptoms, transmission and preventive measures. Majority of the students obtained information about COVID-19 from television, radio, social media, and from family and friends. Overall, the students also demonstrated a positive perception towards COVID-19 mode of transmission and preventive measures.

**Conclusions:** Overall, senior high school students in the Bawku Municipality in Ghana demonstrated an appreciable level of knowledge and positive perception of COVID-19. Students cited television, radio, peer education and social media as their information sources for COVID-19. These media outlets should be prioritized in disseminating COVID 19 information to the public, especially students.

## Background

The novel Coronavirus disease (COVID-19), which emerged in December 2019 in Wuhan, China, is one of the major disease outbreaks the world has ever experienced.^1^ Since the outbreak was first announced in Wuhan in December 2019, more than 48.7 million people have been infected with the virus, with over one million deaths reported worldwide (as of November 6, 2020). ^2,3^ The morbidity and mortality associated with COVID-19 changes rapidly due to the virulent nature of the virus. ^4^ This has led to many healthcare systems around the world struggling to cope with COVID-19 care and treatment, with many patients requiring comprehensive care to survive. ^5^ Global efforts to fight the pandemic has seen many countries adopt COVID-19 preventive measures, including aggressive measures such as border closures and bans on public gatherings. ^6–8^

The incubation period of COVID-19 is usually between 2-14 days and presents common symptoms such as fever, dry cough, feeling of tiredness and shortness of breath. ^9–12^ An individual can be infected with the virus when in close contact with an infected person via respiratory droplets from an infected person. Transmission of the virus may also occur when an individual touches his/her mouth, nose or eyes with the hands contaminated with the virus. ^13,14^ Recent evidence suggests that the virus can be transmitted through aerosols, ^15^ but limited evidence currently exist to fully understand its etiology. Therefore, current strategies to combat the disease are based on the evidence that are available locally and globally.

The first cases of COVID-19 were reported in Ghana on March 12, 2020, ^16^ and as of November 7, 2020, Ghana has recorded a total number of 48,904 cases with 320 deaths. ^3^ In its efforts to curtail the spread of the virus in Ghana, the Ghanaian government instituted a number of policy measures including, border closure and partial lockdowns, testing, contact tracing and treating as well as calling on citizens to observe COVID-19 safety protocols. ^17,18^ Schools and universities were closed down, and only reopened for final year students in the junior and senior high schools, and tertiary institutions in June 2020 to prepare for their final examination. ^19^ Although the Government has, in September 2020, eased restrictions on public gatherings, and reopened borders allowing for international travels to and from the country, all schools and universities have remained closed until January 2021. Before all other students return to schools, especially those at the Senior High Schools who are the active group and likely to be at increased risk of COVID 19, it is important to evaluate their knowledge and perceptions regarding the virus. This is particularly important to help the government and the education authorities to understand what needs to be addressed to prevent the spread of the virus in schools. From the best of our knowledge, no study has yet investigated the knowledge and perception of students about COVID 19 in the country. It is for this reason that this study was designed with the aim to assess the knowledge and perceptions about COVID-19 among Senior High School students.

## Methods

### Study Design and Participants

A descriptive cross-sectional study was conducted using a structured questionnaire to solicit responses from senior high schools’ students aged 18 years and older in Ghana. We recruited students from four senior high schools in the Bawku Municipality in the Upper East Region, Ghana, using a convenient sampling approach. These were the final year students the Government granted to return to take their final examination. Using a 5% margin of error with a 95% confidence interval, and an assumption that 50% of students had knowledge about COVID-19, we arrived at a minimum sample size of 384 respondents. However, we reached out to 631 students to invite them to participate in the study, but 7 students were ineligible. Therefore, a total of 624 students participated in the study by completing our survey questionnaire. The questionnaire collected data on students’ socio-demographics profile, knowledge about COVID-19 symptoms, transmission and preventive/safety measures. To ensure the validity, clarity and relevance of the questionnaire, our survey instrument was pretested among 35 randomly selected students. Ethical approval for the study was obtained from the Christian Health Association (CHAG) Institutional Review Board. Permission was also sought from the schools prior to the data collection.

### Primary variables of interest

The variables of interest included: participants’ knowledge about COVID-19 symptoms, transmission and preventive measures; source of information about COVID-19; and participants’ perception about COVID-19. We assessed whether participants had knowledge of one or more symptoms of COVID-19 or not. Symptoms of COVID-19 that were assessed included; fever, dry cough, tiredness, body aches, diarrhea, sore throat, nasal congestion, conjunctivitis, headache, loss of taste or smell, rash on fingers or toes.^2^

On participants’ knowledge of COVID-19 transmission, we assessed the following: COVID-19 can be transmitted via droplets from the nose or mouth (yes, no); COVID-19 can be transmitted by touching droplets on surfaces or objects (yes, no); COVID-19 asymptomatic person can transmit the virus (yes, no); and COVID-19 can be transmitted among adolescents and young adults (yes, no). Participants knowledge on COVID-19 preventive measures that were also assessed were: handwashing or hand sanitizing can prevent COVID-19 (yes, no); wearing a face mask can prevent COVID-19 (yes, no); social distancing can prevent COVID-19 (yes, no); avoiding crowdedness in public places can prevent COVID-19 (yes, no); and covering the nose and mouth while coughing can prevent COVID-19 (yes, no).

Regarding the medium by which participants heard information about COVID-19, we assessed various sources of information. The sources of information included radio (yes, no), television (yes, no), social media (yes, no), training/educational program (yes, no), family and friends (yes, no), official Government website (yes, no), and newspaper (yes, no).

We also assessed various perceptions about COVID-19. These perceptions included: COVID-19 is transmitted by eating contaminated food (yes, no); COVID-19 is transmitted by drinking contaminated water (yes, no); COVID-19 is transmitted by mosquitos (yes, no); there is no need to take any measures because COVID-19 is not deadly (yes, no); there is no need to take any measures because COVID-19 is not real (yes, no); there is high risk of getting infected with COVID-19 in school (yes, no); the school has put in place adequate COVID 19 protocols (yes, no); and testing of temperature with infrared thermometer at the entrance to the school and classrooms is an adequate screening exercise (yes, no).

### Data analysis

Data analysis was conducted using SAS version 9.3 (SAS Institute, Cary, NC). Descriptive analysis was used to assess participants’ knowledge about COVID-19 symptoms, transmission and preventive measures; as well as the source of information and perceptions about COVID-19.

## Results

The mean age of respondent**s** was 19.9 years ± 1.48, and males constituted the majority in our study population (54.2%). Most of the respondents’ mothers had no formal education (61.7%), neither their fathers (58.0 %). More than a third of the respondents had a health insurance (78.5%) [Table 1].

**Table 1:**
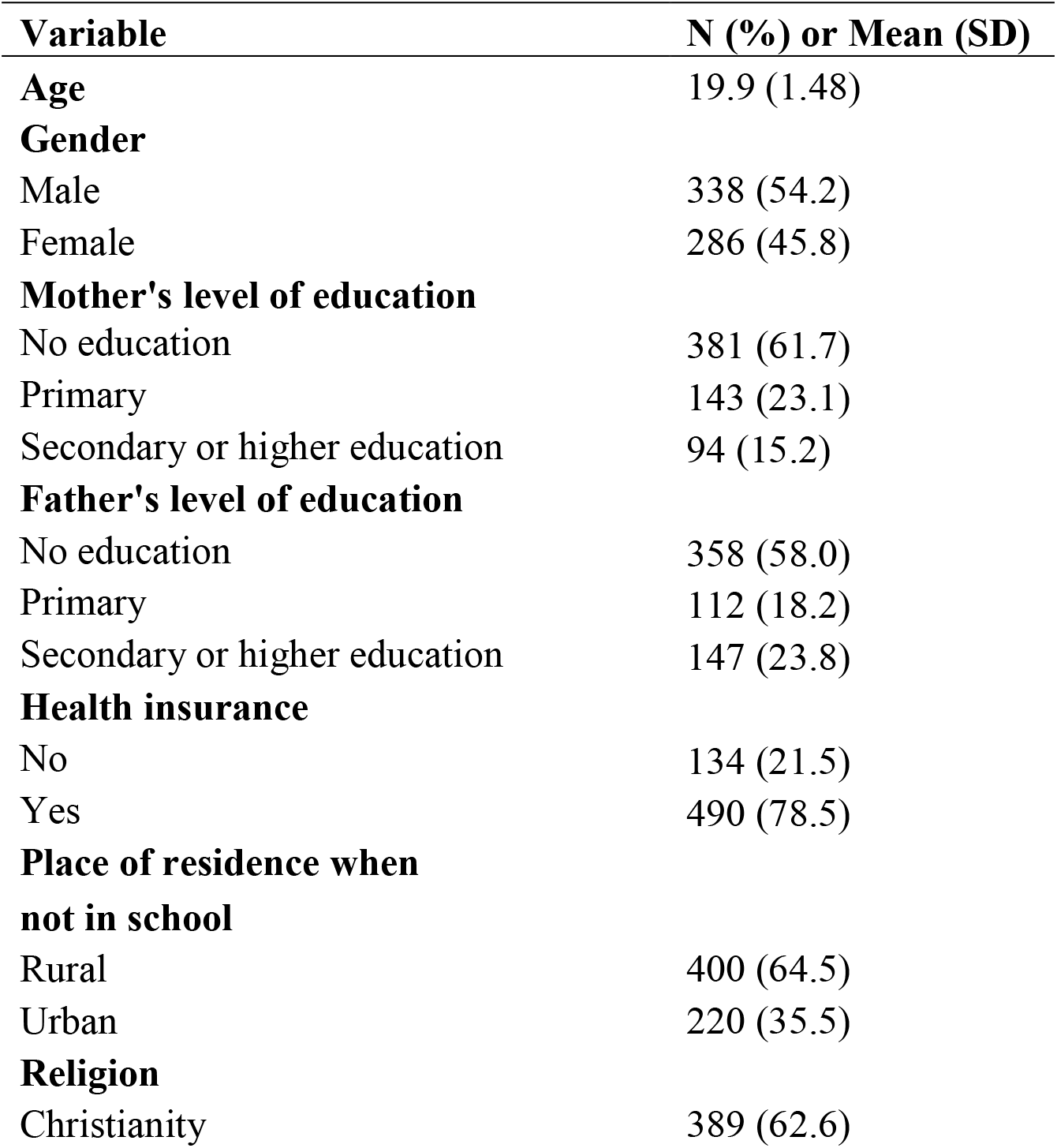
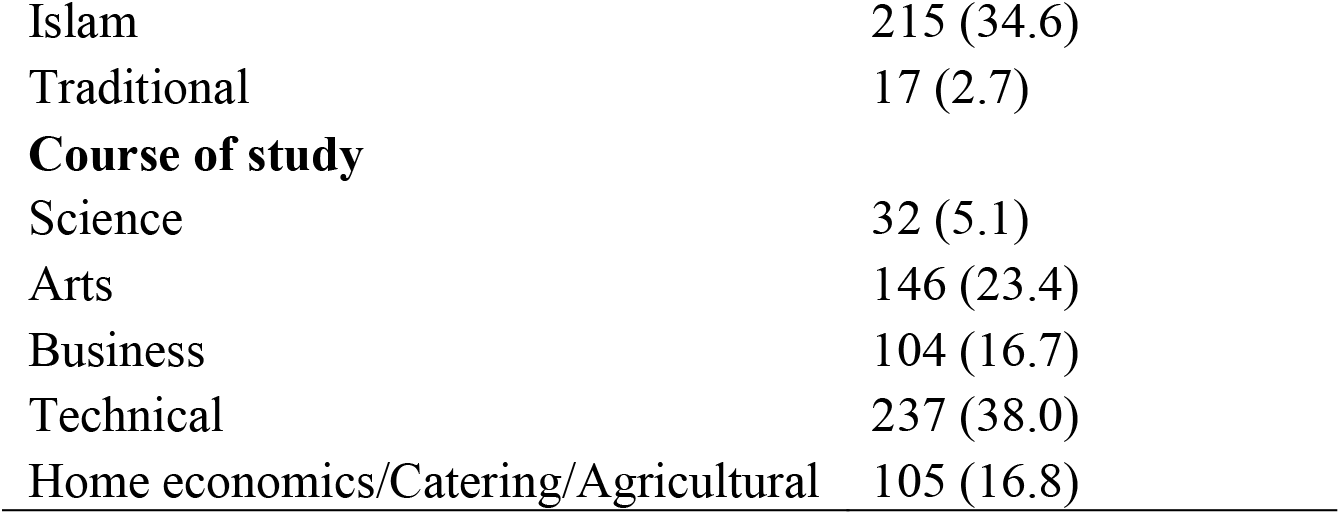
Characteristics of the study sample (n=624)

| **Variable** | **N (%) or Mean (SD)** |
| --- | --- |
| **Age** | 19.9 (1.48) |
| **Gender** |  |
| Male | 338 (54.2) |
| Female | 286 (45.8) |
| **Mother's level of education** |  |
| No education | 381 (61.7) |
| Primary | 143 (23.1) |
| Secondary or higher education | 94 (15.2) |
| **Father's level of education** |  |
| No education | 358 (58.0) |
| Primary | 112 (18.2) |
| Secondary or higher education | 147 (23.8) |
| **Health insurance** |  |
| No | 134 (21.5) |
| Yes | 490 (78.5) |
| **Place of residence when not in school** |  |
| Rural | 400 (64.5) |
| Urban | 220 (35.5) |
| **Religion** |  |
| Christianity | 389 (62.6) |
| Islam | 215 (34.6) |
| Traditional | 17 (2.7) |
| **Course of study** |  |
| Science | 32 (5.1) |
| Arts | 146 (23.4) |
| Business | 104 (16.7) |
| Technical | 237 (38.0) |
| Home economics/Catering/Agricultural | 105 (16.8) |

Almost all (99%) of the respondents reported having knowledge of one or more symptoms of COVID-19. Concerning the knowledge on the transmission of the virus, 92.3% of respondents asserted that COVID-19 can be transmitted via respiratory droplets from the nose or mouth of an infected person. About 94% of respondents reported that Covid-19 can be transmitted by touching droplets on surfaces or objects. Majority (98.9%) of the respondents reported that handwashing or hand sanitizing could prevent COVID-19, while 98.1% revealed that wearing a facemask could also prevent the transmission of COVID-19. Most respondents demonstrated knowledge that social distancing and avoiding crowded places could prevent COVID-19 (98.1%) [Table 2].

**Table 2:** Knowledge about COVID-19 symptoms, transmission and preventive measures

| Variable | Frequency (%) |
| --- | --- |
| Knowledge of COVID-19 symptoms |  |
| None | 6 (1.0) |
| One or more symptoms | 618 (99.0) |
| COVID-19 can be transmitted via droplets from the nose or mouth |  |
| No | 48 (7.7) |
| Yes | 576 (92.3) |
| COVID-19 can be transmitted by touching droplets on surface or objects |  |
| No | 35 (5.6) |
| Yes | 588 (94.4) |
| COVID-19 asymptomatic person can transmit the virus |  |
| No | 111 (17.8) |
| Yes | 513 (82.2) |
| COVID-19 can be transmitted among adolescents and young adults |  |
| No | 141 (22.6) |
| Yes | 483 (77.4) |
| Handwashing or hand sanitizing can prevent COVID-19 |  |
| No | 7 (1.1) |
| Yes | 616 (98.9) |
| Wearing a face mask can prevent COVID-19 |  |
| No | 12 (1.9) |
| Yes | 612 (98.1) |
| Social distancing can prevent COVID-19 |  |
| No | 12 (1.9) |
| Yes | 612 (98.1) |
| Avoiding crowdedness in public places can prevent COVID-19 |  |
| No | 12 (1.9) |
| Yes | 612 (98.1) |
| Covering the nose and mouth while coughing can prevent COVID-19 |  |
| No | 21 (3.4) |
| Yes | 602 (96.6) |
| Avoiding touching of the eyes, nose and mouth can prevent COVID-19 |  |
| No | 51 (8.2) |
| Yes | 573 (91.8) |

More than 90% of participants obtained information about COVID-19 via radio and television. This was followed by social media (89.9%), and family and friends (89.1%) [Table 3]. The least source of information about COVID-19 was via official government websites (56.6%) and newspapers (59.7%).

**Table 3:**
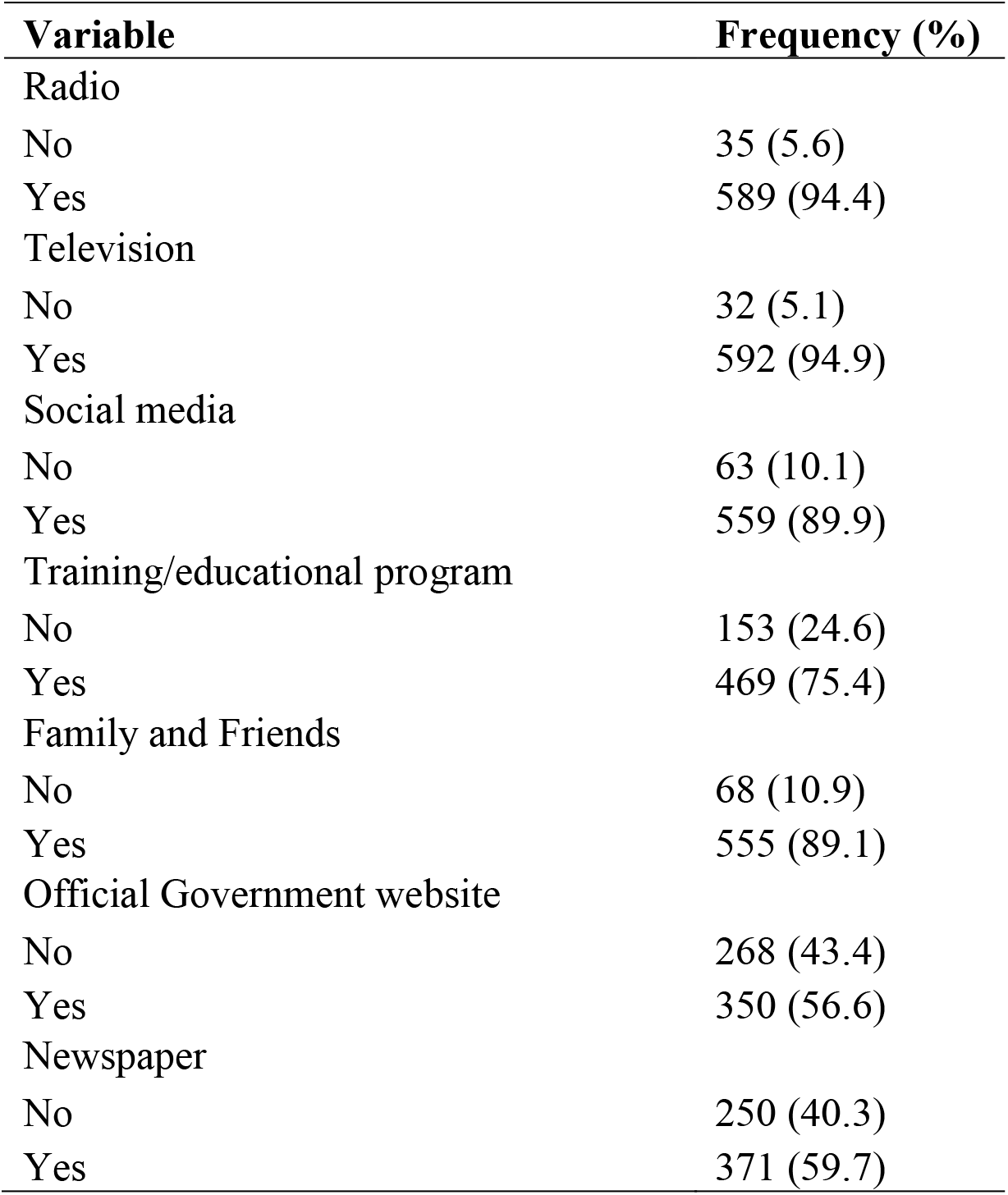
Source of information about COVID-19

| **Variable** | **Frequency (%)** |
| --- | --- |
| Radio |  |
| No | 35 (5.6) |
| Yes | 589 (94.4) |
| Television |  |
| No | 32 (5.1) |
| Yes | 592 (94.9) |
| Social media |  |
| No | 63 (10.1) |
| Yes | 559 (89.9) |
| Training/educational program |  |
| No | 153 (24.6) |
| Yes | 469 (75.4) |
| Family and Friends |  |
| No | 68 (10.9) |
| Yes | 555 (89.1) |
| Official Government website |  |
| No | 268 (43.4) |
| Yes | 350 (56.6) |
| Newspaper |  |
| No | 250 (40.3) |
| Yes | 371 (59.7) |

**Table 4:** Perceptions about COVID-19 among Senior High School students in Ghana

| Variable | Frequency (%) |
| --- | --- |
| COVID-19 is transmitted by eating contaminated food |  |
| No | 450 (72.1) |
| Yes | 174 (27.9) |
| COVID-19 is transmitted by drinking contaminated water |  |
| No | 456 (73.2) |
| Yes | 167 (26.8) |
| COVID-19 is transmitted by mosquitos |  |
| No | 500 (80.3) |
| Yes | 123 (19.7) |
| There is no need to take any measures because COVID-19 is not deadly |  |
| No | 498 (79.8) |
| Yes | 126 (20.2) |
| There is no need to take any measures because COVID-19 is not real |  |
| No | 517 (82.9) |
| Yes | 107 (17.1) |
| There is high risk of getting infected with COVID-19 in school |  |
| No | 169 (27.1) |
| Yes | 455 (72.9) |
| The school has put in place adequate COVID 19 protocols |  |
| No | 87 (13.9) |
| Yes | 537 (86.1) |
| Testing of temperature with infrared thermometer at the entrance to the school and classrooms is an adequate screening exercise |  |
| No | 83 (13.3) |
| Yes | 540 (86.7) |

Participants who perceived that COVID-19 is transmitted by eating contaminated food and drinking contaminated water constituted 27.9% and 26.8%respectively. Some participants perceived that COVID-19 is transmitted by mosquitos (19.7%). The participants who perceived that there is no need to take any measures because COVID-19 is not deadly and not real, constituted 20.2% and 17.1% respectively of the sample.

## Discussions

This was a description cross sectional study to explore the understanding and perception of senior high school students designed in the Upper East Region regarding the COVID-19 pandemic. The findings revealed that Senior High School students in the Bawku municipality in Ghana are aware of the COVID-19 symptoms, mode of transmission and the preventive measures. Sources of COVID-19 information that were cited by many of the students include television, radio, social media, and from family and friends. Majority of the students had a positive perception towards the COVID-19 measures put in place to prevent the spread of the disease.

The results suggest that almost all the respondents (99%) had knowledge on one or more symptoms about COVID-19. This finding is in line with other studies ^20, 21^ where more than 78% of respondents were able to identify some of the symptoms of the virus. Haven enough knowledge on the symptoms of COVID 19 could help in the early detection of the pandemic and seeking early treatment. With regards on how the virus is transmitted, more than two thirds of the respondents demonstrated good knowledge. This is in contrast with a study carried out by Kushalkumar et al, were a lesser number of respondents knew the mode of COVID 19 transmission. ^22^ Respondents in our study (91%) also demonstrated good knowledge on the prevention methods of COVID 19. This finding collaborates with what Zhong et al studied in China in assessing the public knowledge, attitudes, practice of COVID 19, and Taghrir et al study in Iran, which investigated medical students’ knowledge, prevention behavior and risk perception of COVID 19. ^23, 24^ The divergence in the findings reported by these studies and our study could be that whilst our study only focused on Senior High School students, the previous two studies focused on the general population, and medical students. However, some other studies have findings that are similar to our findings, ^25,26,23^ although the respondents in these three studies included health workers, medical and non-medical students and the general population. A recent review of evidence by Puspitasari and colleagues from studies conducted in the United States, United Kingdom, Italy, Jordan and China concluded that information about COVID pandemic was well received by the general public as the majority of the studies reported high level of knowledge about disease. ^27^

The knowledge of COVID-19 symptoms, transmission and preventive measures demonstrated among the students surveyed might be an indication that messages on COVID-19 being disseminated through the various electronic media by both the Government and its partner organizations could be having a positive impact on students’ knowledge of COVID-19. The President of the Republic, through his monthly update on the COVID situation, persistently called on citizens and people living in Ghana particularly schools to observe the measures/protocols being implemented by the Government to combat the pandemic. This might have led to the students increased knowledge of the COVID-19 symptoms, transmission and preventive measures. One other possible explanation might be that the students had increased access to available information above COVID-19. Prior to the final year students returning to school to complete their final exams, we argue that they were exposed, and had increased access to information regarding COVID-19, including information disseminated through television news, radio and social media. This increased access to information might have informed their appreciable knowledge, and positive perception of the COVID-19 prevention measures. Following the COVID-19 pandemic, the Ministry of Health and the Ghana Health Service have used television, radio and the social media to educate the citizenry about the virus, and this has informed our assumption and argument of increased access of COVID-19 information among the students. This finding is consistent with some other studies where the media including social media have been identified as the main source of information on COVID-19. ^28, 29^

Our findings also emphasize the important role peers or family members play in the dissemination of COVID-19 information. Our results show that, apart from the electronic and social media, students received information about COVID-19 from family members and friends. Our results however suggest that that less than two thirds of the students obtained information on the pandemic via official government websites and newspapers. This may be telling us that Ghanaian students at the Senior High Schools do not patronize the Ghanaian print media, nor care to access government websites from important health information. The poor patronage of government websites among students could be attributed to the poor internet connectivity in the country and high cost of internet service. In contrast increase access of information from electronic and social media could be largely attributed to the availability of numerous radio and TV stations and channels as well as the wide coverage of these stations.^30^ Arguably, this has made access to different types of information among the general public easier. This argument is supported by findings from one study conducted in northern Ghana, suggesting that radio and television were the commonest channel through which most people accessed information about COVID-19.^30^ Elsewhere in Pakistan, Mubeen reported that most young adults accessed information about the virus from TV, radio and social media.^31^

In terms of what the students perceived about the COVID-19 crisis, majority of the students in our study had a positive perception about COVID-19 transmission and the measures in place to combat it. This positive perception could be attributed to the appreciable level of knowledge on the COVID-19 discussed earlier on. However, about one fifth of students perceived that COVID-19 was not deadly while others perceived that the virus was not real. This is a major concern as such students are not likely to fully observe the COVID-19 safety protocols at school. This puts them and their colleagues at increased risk of acquiring the virus. These findings aligned well with what Saba et al reported in their study in northern Ghana where participants perceived that the pandemic was sent by God to punish mankind for their sins.^30^ These findings highlights the need to intensify education in the northern part of Ghana about the diseases.

Our study had a number of limitations. There is a likelihood of recall bias as the responses provided by the students were self-reported. Further we want to caution readers for any attempt to generalize the findings of the study to the general senior high schools’ students’ population in the country as our sample is not representative of the entire country. Despite these limitations, the finding provides useful information about students’ knowledge and perceptions about COVID-19 and could be useful to local authorities, particularly the study setting to make decisions to stop the spread of the disease among students when schools are finally reopened.

## Conclusion

In this study, students of senior high school demonstrated good knowledge about COVID-19 symptoms and transmission and had a positive perception about the measures put in place to tackle the pandemic in Ghana. The results show that students generally obtained information on COVID-19 from television, radio, social media and family and friends. Moving forward, and in the future, the authorities should consider using these media as a platform to drum home their message of diseases prevention.

## Data Availability

All data referred to in this manuscript are form it.

## Acknowledgements

We wish to express our appreciation to all the study participants and schools for participating in this study.

## Funding Information

Self-funded

## Competing Interests

The authors declare that they have no competing interests.

## Notes

### Competing Interest Statement

The authors have declared no competing interest.

### Funding Statement

This study is self funded.

### Author Declarations

Christian Health Association of Ghana Research Unit

## References

1. World Health Organization. Novel Coronarius - Coronavirus Disease 2019. https://www.who.int/docs/default-source/coronaviruse/situation-reports/20200423-sitrep-94-covid-19.pdf. Published 2020. Accessed September 13, 2020.

2. World Health Organization. WHO Director-General’s opening remarks at the media briefing on COVID-19 - 11 March 2020. https://www.who.int/dg/speeches/detail/who-director-general-s-opening-remarks-at-the-media-briefing-on-covid-1911-march-2020. Published 2020. Accessed September 13, 2020.

3. Johns Hopkins University. Coronavirus resource Center. https://coronavirus.jhu.edu/map.html. xPublished 2020. Accessed September 13, 2020.

4. Fisher D, Heymann D. Q&A: The novel coronavirus outbreak causing COVID-19. BMC Med. 2020;18:57. doi:10.1186/s12916-020-01533-w

5. Abate SM, Ali SA, Mantfardo B, Basu B. Rate of intensive care unit admission and outcomes among patients with coronavirus: A systematic review and Meta-analysis. PLoS One. 2020;15(7):e0235653. doi:10.1371/journal.pone.0235653

6. World Bank. Assessing the Economic Impact of Covid-19 and Policy Responses in Sub-Saharan Africa.; 2020. https://elibrary.acbfpact.org/cgi-bin/acbf?e=d-010off-acbf--00-1--0---0-10-TX-1%2C1%2C1%2C1-4-------0-11l--11-en-50---20-home---01-3-1-0000-0-11-0utfZz8-00&a=d&d=HASHd1df5b364a9ea7b007194b&gg=0.

7. World Health Organization. Coronavirus Disease 2019 (COVID-19) Situation Reports - 72. https://www.who.int/docs/default-source/coronaviruse/situation-reports/20200324-sitrep-64-covid-19.pdf?sfvrsn=703b2c40_2%0Ahttps://www.who.int/docs/default-source/coronaviruse/situation-reports/20200401-sitrep-72-covid-19.pdf?sfvrsn=3dd8971b_2. Published 2020. Accessed September 24, 2020.

8. Cirrincione L, Plescia F, Ledda C, et al. COVID-19 Pandemic: Prevention and protection measures to be adopted at the workplace. Sustain. 2020;12:3603. doi:10.3390/SU12093603

9. Guan W, Ni Z, Hu Y, et al. Clinical characteristics of coronavirus disease 2019 in China. N Engl J Med. 2020;382(18):1708–1720. doi:10.1056/NEJMoa2002032

10. Huang C, Wang Y, Li X, et al. Clinical features of patients infected with 2019 novel coronavirus in Wuhan, China. Lancet. 2020;395(10223):497–506. doi:10.1016/S0140-6736(20)30183-5

11. Nanshan C, Min Z, Xuan D, et al. Epidemiological and Clinical Characteristics of 99 Cases of 2019 Novel Coronavirus Pneumonia in Wuhan, China: A Descriptive Study. Lancet. 2020;395(10223):507–513. doi:10.1016/S0140-6736(20)30211-7

12. Zhao X-Y, Xu X-X, Yin H-S, et al. Clinical Characteristics of Patients with 2019 Coronavirus disease in a non-Wuhan area of Hubei Province, China: a retrospective study. BMC Infect Dis. 2020;20:311. doi:10.21203/rs.3.rs-15734/v1

13. World Health Organization. Modes of transmission of virus causing COVID-19: implications for IPC precaution recommendations: Scientific brief. https://www.who.int/docs/default-source/coronaviruse/20200329-scientific-brief1-final-corrected2-revised-final.pdf?sfvrsn=cb817b3a_6&ownload=true. Published 2020. Accessed September 13, 2020.

14. Yan-Rong G, Qing-Dong C, Zhong-Si H, et al. The origin, transmission and clinical therapies on coronavirus disease 2019 (COVID-19) outbreak – an update on the status. BMC Mil Med Res. 2020;7:11. doi:10.1186/s40779-020-00240-0

15. Jayaweera M, Perera H, Gunawardana B, Manatunge J. Transmission of COVID-19 virus by droplets and aerosols?: A critical review on the unresolved dichotomy. Environ Res. 2020;188(2020):109819. doi:10.1016/j.envres.2020.109819

16. Ghana Health Service. Ghana Confirms Two cases of COVID-19. http://ghanahealthservice.org/covid19/downloads/covid_19_first_confirmed_GH.pdf. Published 2020. Accessed September 13, 2020.

17. Ghana Statistical Srvice. Mobility analysis to support the Government of Ghana in responding to the COVID-19 outbreak. https://statsghana.gov.gh/COVID-19pressreleasereport-analysisoverview-final.pdf. Published 2020. Accessed September 13, 2020.

18. Sarfo AK, Karuppannan S. Application of Geospatial Technologies in the COVID-19 Fight of Ghana. Trans Indian Natl Acad Eng. 2020;5(2):193–204. doi:10.1007/s41403-020-00145-3

19. Graphic Online. Schools and universities to reopen for final year students. https://www.graphic.com.gh/news/politics/schools-and-universities-to-reopen-for-final-year-students.html. Published 2020. Accessed September 24, 2020.

20. Backer J, Klinkenberg D, Wallinga J. (2020). Incubation period of 2019 novel coronavirus (2019-nCoV) infections among travellers from Wuhan, China, 20–28 January 2020. Euro Surveill. 25(5):2000062. https://doi.org/10.2807/1560-7917.ES.2020.25. 5.2000062.

21. Abdelhafiz AS, Mohammed Z, Ibrahim ME, et al. (2020) Knowledge, perceptions, and attitude of Egyptians towards the novel coronavirus disease (COVID-19). J Community Health. 45:881–890. https://doi.org/10.1007/s10900-020-00827-7.

22. Kushalkumar H. Gohel, et al., (2020). Clinical Epidemiology and Global Health, https://doi.org/10.1016/j.cegh.

23. Zhong B, Luo W, Li H, et al (2020). Knowledge, attitudes, and practices towards COVID-19 among Chinese residents during the rapid rise period of the COVID-19 outbreak: a quick online cross-sectional survey. Int J Biol Sci., 16(10):1745–1752. doi:10.7150/ijbs.45221.

24. Taghrir MH, Borazjani R, Shiraly R. COVID-19 and Iranian medical students; A survey on their related-knowledge, preventive behaviors and risk perception. Arch Iran Med. 2020;23(4):249–254. https://doi.org/10.34172/aim.2020.06

25. Zhang M, Zhou M, Tang F, et al (2020). Knowledge, attitude and practice regarding COVID-19 among health care workers in Henan, China. J Hosp Infect.105(2):183–187. doi:10.1016/j.jhin.2020.04.012.

26. Alzoubi H, Alnawaiseh N, Lubad MA, Aqel A, Al-Shagahin H. (2020). COVID-19 - knowledge, attitude and practice among medical and non-medical university students in Jordan. J Pure Appl Microbiol.14(1):17–24. doi:10.22207/JPAM.14.1.04

27. Puspitasari et al (2020). Knowledge, Attitude, and Practice During the COVID-19 Pandemic: A Review. Journal of Multidisciplinary Healthcare.13 727–733.

28. Giao, H., Han, N. T. N., Van Khanh, T., et al (2020). Knowledge and attitude toward COVID-19 among healthcare workers at District 2 Hospital, Ho Chi Minh City. Asian Pacifc J Trop Med, 13, 3–5. https://doi.org/10.4103/1995-7645.280396.

29. Bhagavathula, A. S., Wafa Ali Aldhaleei, W.A., Rahmani, J., et al (2020). Knowledge and perceptions of COVID-19 among health care workers: Cross-sectional study. MIR Public Health Surveillance, 6(2), e19160. https://doi.org/10.2196/19160.

30. Saba C.K.S, Karikari A. B, et al (2020). COVID-19: Knowledge, perceptions and attitudes of residents in the Northern Region of Ghana, West Africa. Resaerchgate. doi:10.20944/preprints202008.0060.v1

31. Mubeen, S., Kamal, S., Kamal, S., and Balkhi, F. (2020). “Knowledge and Awareness Regarding Spread and Prevention of COVID-19 among the Young Adults of Karachi.” Journal of the Pakistan Medical Association 1. doi:10.5455/JPMA.40

